# Indepth characterization of a cohort of individuals with missense and loss-of-function variants disrupting *FOXP2*

**DOI:** 10.1101/2022.06.01.22275851

**Authors:** Lottie Morison, Elisabeth Meffert, Miriam Stampfer, Irene Steiner-Wilke, Brigitte Vollmer, Katrin Schulze, Tracy Briggs, Ruth Braden, Adam P. Vogel, Daisy Thompson-Lake, Chirag Patel, Edward Blair, Himanshu Goel, Samantha Turner, Ute Moog, Angelika Riess, Frederique Liegeois, David A. Koolen, David J. Amor, Tjitske Kleefstra, Simon E. Fisher, Christiane Zweier, Angela T. Morgan

## Abstract

**Background:** Heterozygous disruptions of *FOXP2* were the first identified molecular cause for severe speech disorder; childhood apraxia of speech (CAS), yet few cases have been reported, limiting knowledge of the condition.

**Methods:** Here we phenotyped 29 individuals from 18 families with pathogenic *FOXP2*-only variants (13 loss-of-function, 5 missense variants; 14 males; aged 2 years to 62 years). Health and development (cognitive, motor, social domains) was examined, including speech and language outcomes with the first cross-linguistic analysis of English and German.

**Results:** Speech disorders were prevalent (24/26, 92%) and CAS was most common (23/26, 89%), with similar speech presentations across English and German. Speech was still impaired in adulthood and some speech sounds (e.g. ‘th’, ‘r’, ‘ch’, ‘j’) were never acquired. Language impairments (22/26, 85%) ranged from mild to severe. Comorbidities included feeding difficulties in infancy (10/27, 37%), fine (14/27, 52%) and gross (14/27, 52%) motor impairment, anxiety (6/28, 21%), depression (7/28, 25%), and sleep disturbance (11/15, 44%). Physical features were common (23/28, 82%) but with no consistent pattern. Cognition ranged from average to mildly impaired, and was incongruent with language ability; for example, seven participants with severe language disorder had average non-verbal cognition.

**Conclusions:** Although we identify increased prevalence of conditions like anxiety, depression and sleep disturbance, we confirm that the consequences of *FOXP2* dysfunction remain relatively specific to speech disorder, as compared to other recently identified monogenic conditions associated with CAS. Thus, our findings reinforce that *FOXP2* provides a valuable entrypoint for examining the neurobiological bases of speech disorder.

**What is already known on this topic:** Heterozygous disruptions of *FOXP2* were the first identified molecular cause for severe speech disorder; childhood apraxia of speech (CAS), yet few cases have been reported, limiting knowledge of the condition.

**What this study adds:** Here we provide the most comprehensive characterisation of individuals with pathogenic *FOXP2* variants, almost doubling the number of published families to date. We provide the first cross-linguistic analysis of speech and language across German and English. We show that the phenotype for pathogenic *FOXP2* variants remains relatively specific to speech disorder, compared to phenotypes associated with other monogenic conditions involving CAS.

**How this study might affect research, practice or policy:** This study guides identification of cases with a *FOXP2*-related disorder for a clinical genetic diagnosis, will improve prognostic counselling and lead to better targeted clinical management.

## Introduction

*FOXP2* was the first gene implicated in a developmental speech and language disorder in the absence of intellectual disability (1). A private heterozygous missense variant in *FOXP2* was found to co-segregate with childhood apraxia of speech (CAS) in 15 members of the multi-generational British ‘KE family’, while being absent from all unaffected relatives and from healthy controls (1). The study also characterized a balanced chromosomal translocation with a 7q31.2 breakpoint disrupting *FOXP2* in an unrelated proband with similar speech deficits (1). CAS is a disorder of speech motor planning and programming that manifests in impaired sequencing of speech sounds into syllables and words with the correct prosody (2). The condition is associated with delayed and protracted speech development.

Pathogenic single-nucleotide variants (SNVs) and intragenic indels that disrupt *FOXP2* are rare. To our knowledge, there have been a dozen of these SNVs/indels reported in the literature to date: the original missense variant in the large KE family (1), a nonsense (stop-gain) variant in two siblings and their mother (3); a frameshift in a sporadic patient (4), and eight variants across eight small families (intragenic deletions, nonsense, missense and frameshift variants) (5) (Supplementary Table 1), each occurring in a heterozygous state. The limited number of cases reported may in part be due to the relatively mild speech-focused phenotype associated with pathogenic SNVs/indels of *FOXP2*, compared to other neurogenetic childhood disorders. Whilst debilitating for affected probands and families, a CAS phenotype does not often lead to clinical genetic testing, nor to ascertainment in gene discovery cohorts for other neurodevelopmental disorders such as autism spectrum disorder (ASD) or intellectual disability. All probands with pathogenic *FOXP2* variants reported to date share a severe speech disorder presentation, most commonly CAS. Yet there is preliminary evidence that, in some cases, SNVs or indels of *FOXP2* may cause a broader phenotype including subtle dysmorphology and co-occurring neurodevelopmental features such as ASD (5).

There are other variants that affect *FOXP2* that are not SNVs or indels. A large deletion downstream of *FOXP2* was hypothesised to have a position effect on expression (6). There have also been case series of large heterozygous 7q31 deletions or reciprocal balanced translocations associated with more complex phenotypes involving disruptions of *FOXP2* in addition to neighbouring genes (7-15). Such phenotypes are now clinically defined as having a *FOXP2*-plus related disorder (16).

A systematic prospective cohort study of the phenotype(s) associated with *FOXP2* variants is warranted to guide better identification of cases for clinical genetic diagnosis, improve prognostic counselling, and lead to better targeted intervention. Here we examine speech, language, health, and broader developmental phenotypes associated with pathogenic *FOXP2* SNVs/indels in a cohort of 29 probands from 18 unrelated families [7 previously published but not deeply characterised for speech and language (3-5), 11 novel] expanding the genetic and clinical spectrum of the disorder. For the first time in any phenotypic study of *FOXP2*, the specificity of a linguistic phenotype, relative to broader neurodevelopmental skills (e.g. communication ability compared to domains of social, motor and daily living skills) was examined using standardised tests. A novel cross-linguistic comparison of speech diagnoses in German-versus English-speaking participants was also conducted, to determine homogeneity of the speech phenotype across languages.

## Methods

### Participants

Inclusion criteria was a molecular diagnosis of pathogenic variants (SNVs or intragenic deletions/duplications) in *FOXP2*, in individuals aged ≥6 months. Participants were recruited via clinical genetics colleagues or family self-referral in the Netherlands, France, Britain, Germany, the United States, the United Kingdom and Australia. Ethics approval was obtained from the Royal Children’s Hospital, Melbourne, Human Research Ethics Committee (HREC 37353A). Adult participants and caregivers of child participants provided informed consent to participate in the study. The assessment battery was tailored to cover a wide range of ages and languages.

### Health and development

Health and medical information, including developmental milestones and existing diagnoses of neurodevelopmental conditions, was collected via an established direct (adult) or caregiver survey (17-19). Health professional reports and consults confirmed caregiver survey responses. Feeding [Child Oral and Motor Proficiency Scale [ChOMPS (20)] and drooling [Drooling Impact Scale (DIS) (21)] measures were collected where age appropriate.

### Speech

In English-speaking participants, phonology and articulation were assessed using standardised tools [Diagnostic Evaluation or Articulation and Phonology [DEAP (22)] or Goldman Fristoe Test of Articulation - Second Edition (23). Phonological delay versus disorder was delineated. Both for English (LM, AM) and German-speaking (EM) participants, phonological and articulation errors were also analysed from phonetic transcription of a five minute conversational speech sample. Across both languages, CAS features were rated across three core diagnostic criteria (17, 18, 24): inconsistent speech errors, lengthened and disrupted coarticulatory transitions, and impaired prosody. These three criteria were further operationalised into rateable speech errors [see Supplementary Table 2]. Similarly, dysarthria was assessed using the Mayo Clinic Dysarthria Classification System rating scale (25), and evaluating oral motor structure and function (26). The Intelligibility in Context Scale (ICS) (27) documented how well an individual was understood by conversational partners, with a five point scale ranging from ‘never’ to ‘always’ understood.

### Language and literacy

Receptive vocabulary and expressive vocabulary were assessed using clinician administered standardised tools dependent on an individual’s age and language [see Table 1 for assessments]. Likewise, caregiver administered standardised tools were used to measure speech and language skills. Assessment tools used were dependent on an individuals communicative ability, age and language (Table 1). Literacy abilities were documented by direct (adult) or caregiver report, the Vineland Adaptive Behaviour Scales – Third Edition (28) written communication subdomain, and academic records.

**Table 1.**
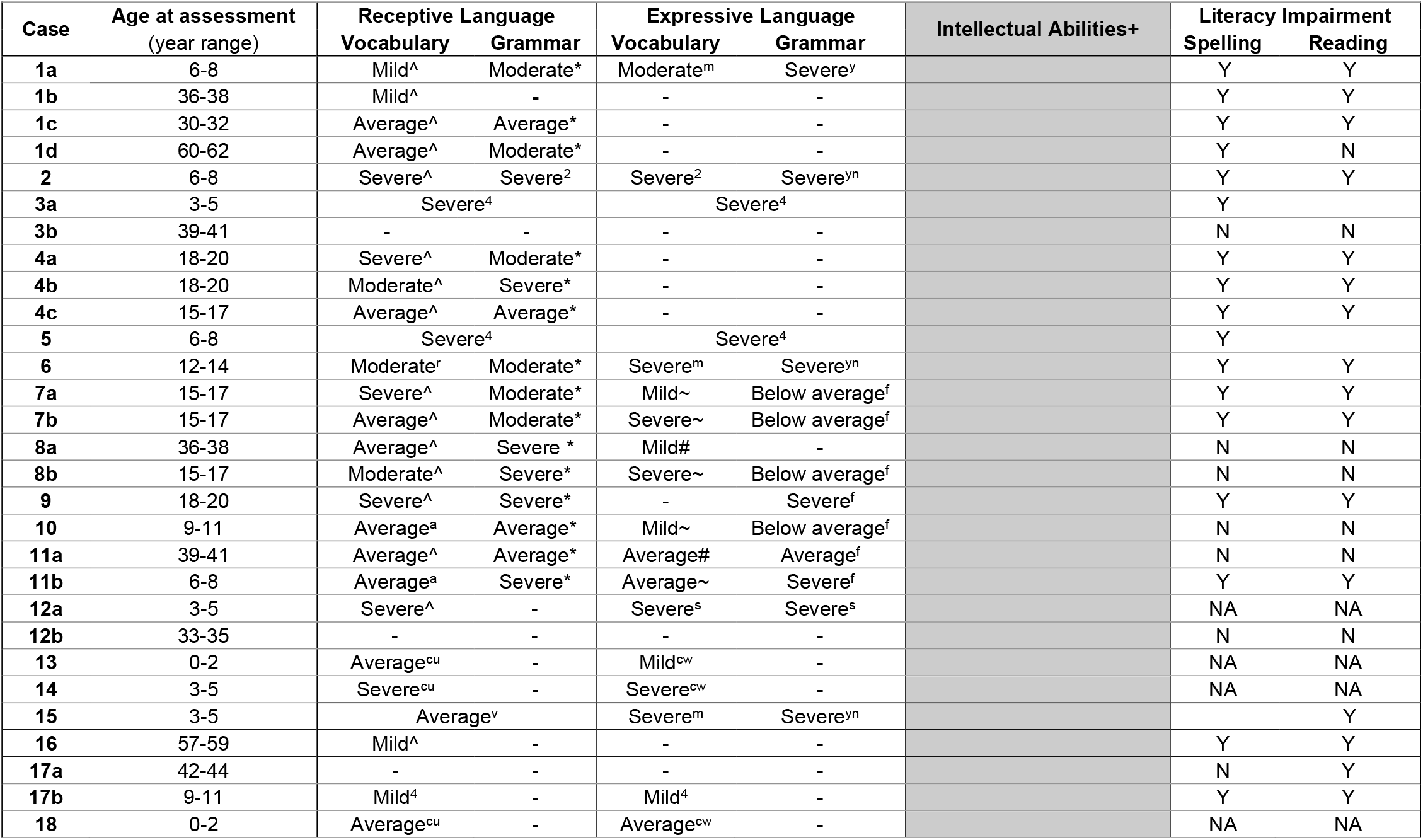
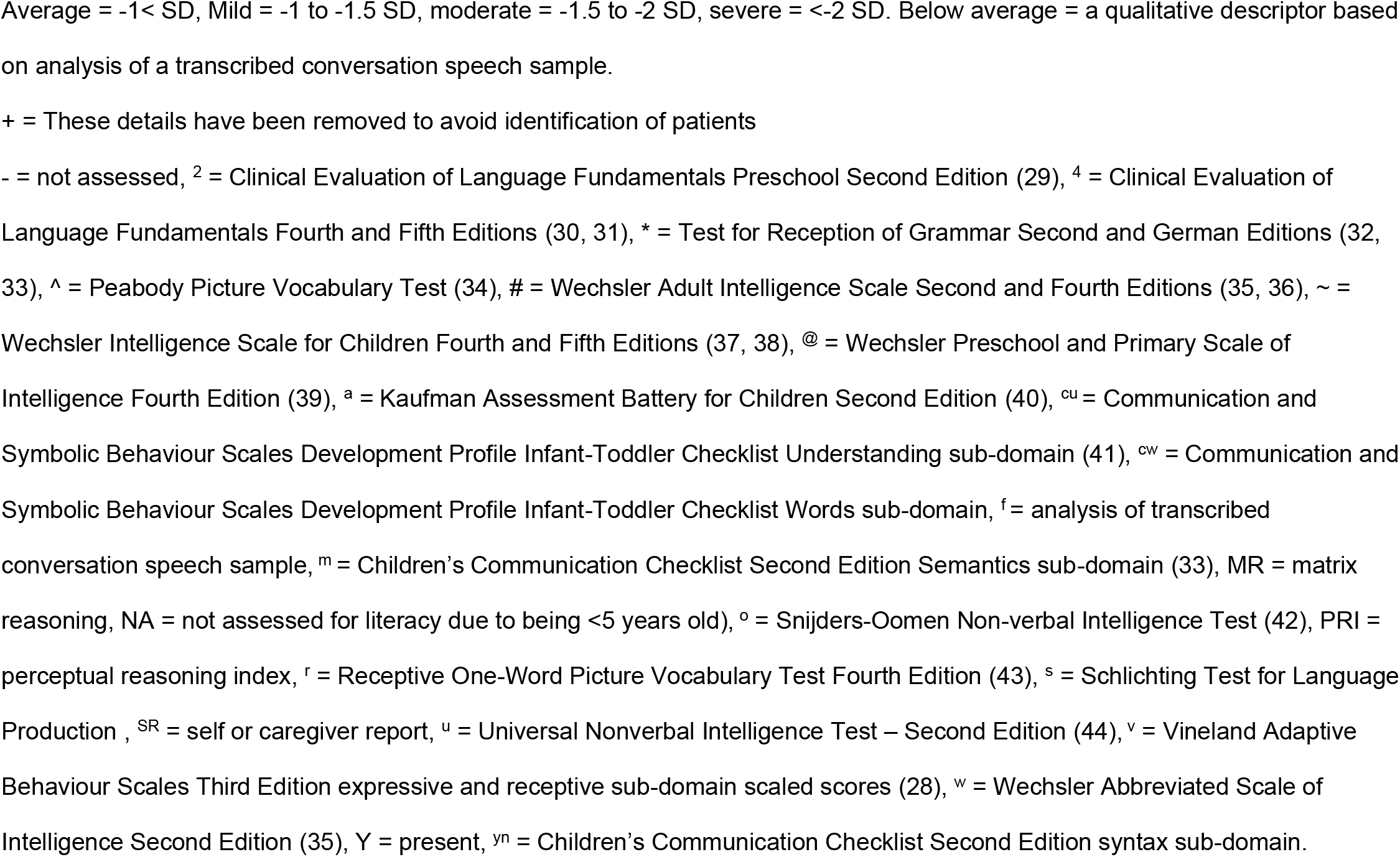
Language skills and cognition in participants with pathogenic missense/loss-of-function variants disrupting *FOXP2*

### Adaptive behaviour and intellectual ability

The VABS-3 provided scores across language, socialisation, self-care, daily living, motor skills, and a composite total adaptive behaviour score (28). A non-parametric Kruskal-Wallis test examined the relative involvement of VABS-3 subdomain scores, and a Wilcoxon Signed Rank test was performed between VABS-3 receptive and expressive language scores, to highlight any differences between these domains.

General intellectual abilities were assessed with the age appropriate Wechsler assessment [see Table 1 for assessments]. Where Full Scale Intelligence Quotient (FSIQ) could not be obtained, Perceptual Reasoning Indexes and Matrix Reasoning subtest scores were calculated from the relevant Wechsler assessment. A further three assessment tools were used to assess intellectual abilities in four children (Table 1). Diagnoses of neurodevelopmental disorders (e.g., autism) were identified by caregiver report, and confirmed with clinical records.

## Results

### Participants

Twenty-nine participants with pathogenic *FOXP2* variants were recruited from 18 families, comprising 11 unreported families [families 1-3, 6, 12-18] and 7 that were previously reported but not deeply characterised for speech and language abilities [families 4, 5, 7-11]. Participants had a median age of 16 years 8 months, range 2 years 7 months to 62 years 7 months, and 14 (48%) were male (Table 2). Most participants were referred for genetic testing due to the proband’s striking speech and language impairment, with the exception of Family 3 (n=2) and participant 18. Family 3 underwent genetic testing as part of research testing for preterm children (ID 3a), and subsequently a variant was also identified in the father (ID 3b). Participant 18 was referred due to microcephaly and dysmorphic facial features.

**Table 2** Genotypes and phenotypes of this cohort

+ = This table has been removed in pre-print to avoid identification of patients

### Genetic results

The 18 families each had a unique *FOXP2* variant, comprising 13 loss of function and 5 missense variants (Table 2). The 13 loss-of-function variants included 3 frameshifts, 4 stop-gain/nonsense variants, 1 splice-site variant, 1 variant abolishing the translational start-site, 3 intragenic deletions, and 1 intragenic duplication. All missense variants were located in the forkhead-box DNA-binding domain of the encoded protein (Figure 1). Participant 16 also had a variant in *PTPN11* (c. 1492 C>T, p.Arg498Trp, likely pathogenic), but did not show any signs of Noonan syndrome, the disorder associated with disruption of that gene (45). Eleven of the 29 participants had confirmed *de novo* variants, 12 inherited their variant from a parent, and for 6 participants the inheritance status was unknown (Table 2, Supplementary Figure 1). Participant 9 was previously reported to have inherited the *FOXP2* variant from their father who had the same variant in a mosaic state, and who was unavailable to take part in the present study. Deletions, sequence variants and phenotypic data were submitted to Decipher (https://decipher.sanger.ac.uk/).

**Figure 1.**
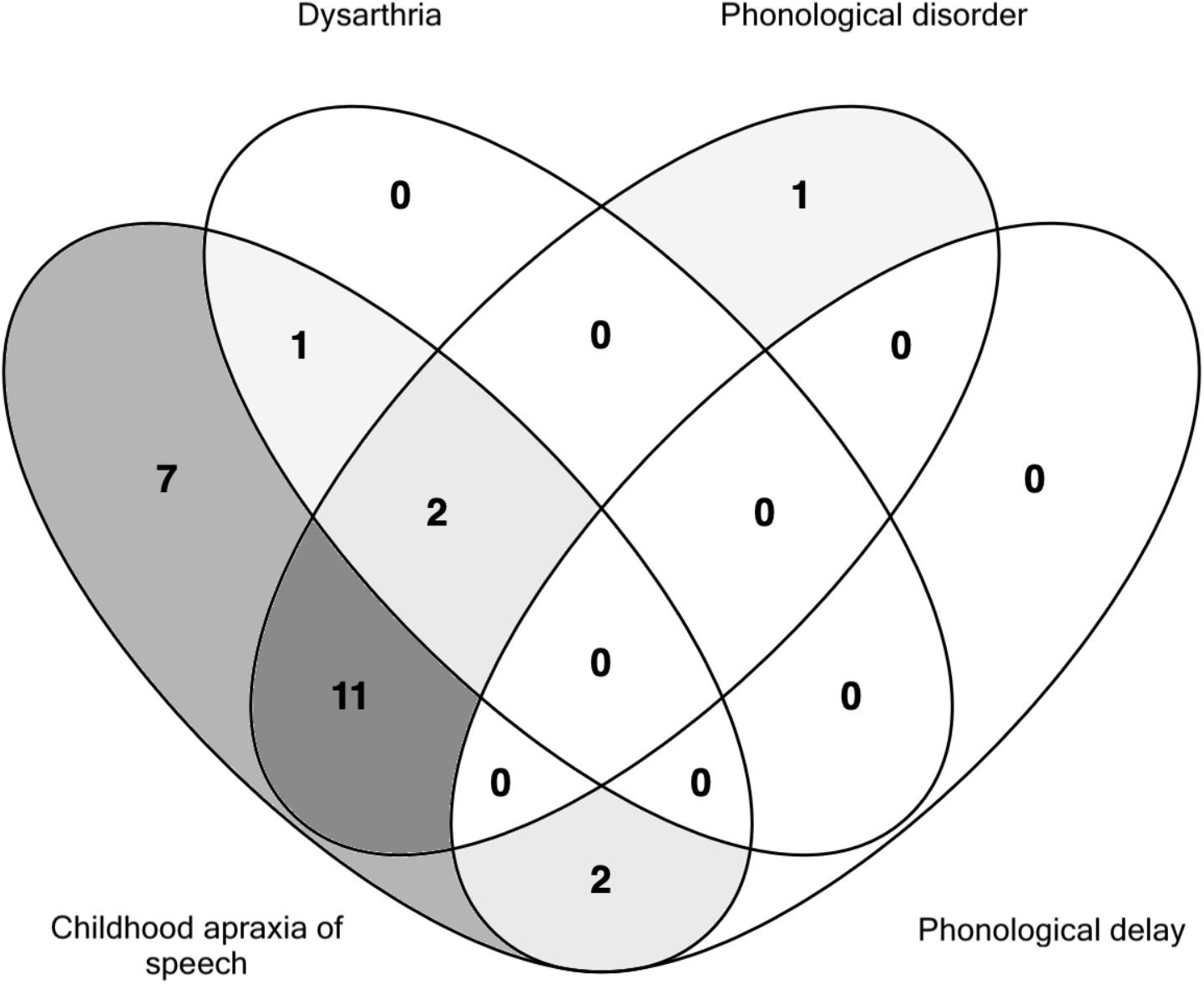
Schematic of 18 pathogenic *FOXP2* variants in this cohort from 29 individuals in 18 families (NM_014491). This figure has been removed to avoid identification of patients

### Health and development

Over a third of assessed participants had feeding difficulties in infancy (10/27), and some had excessive drooling (5/21) in the early years of life (Table 2). Gross motor impairments during early development (14/27) and fine motor impairment (14/27) were also present, with a subset of participants having both fine and gross motor impairment (8/27). Of those with fine motor impairment, two (IDs 12b, 16) had a formal diagnosis of Developmental Coordination Disorder. In a small number of individuals, hypotonia (IDs 6, 12a) or microcephaly (ID 18) was also noted. Three participants had hearing impairment; mild, conductive hearing loss (25-39dBHL, ID 15), moderate mixed hearing loss (40-69dBHL, ID 1d) and severe mixed hearing loss (70-80dBHL, ID 16). Sleep disturbances were relatively common (11/25), mostly characterised as difficulty falling asleep (5/25) or frequent waking (5/25). Visual impairments (8/27) were present (Supplementary Table 3). Physical features were reported in most participants (23/28, four previously reported, Supplementary Table 3), but with no distinct morphological profile across families. Recorded physical features features involved the nose (upturned nose: ID 1a; prominent nose: IDs 1b, 1c; hypoplastic alae nasae: ID 4b, 4c; high nasal root: ID 8b; rounded, fleshy or prominent nasal tip: IDs 1b, 1c, 3a, 5, 8b), philtrum (short/flat philtrum: IDs 2, 18), ears (prominent/protruding ears: IDs 1a, 1b, 12b; anteverted ears: ID 14), eyes (periorbital fullness: IDs 3a, 5, 13; prominent eyes: ID 17b), jaw (retrognathia: IDs 1a, 1b, 3a, 13) and lips (full lips IDs: 1a, 1b, 1c, 4a, 4b, 13, 16; thin upper lip: 3a). In individual cases, mild finger pads (ID 10), tapering fingers (ID 8b), single palmar crease (ID 18), and clinodactyly (ID 12a) were also noted.

#### Co-occuring diagnoses

A quarter of participants had a diagnosis of ASD (7/28; 2 diagnosed in adulthood) and one had autistic traits but did not meet criteria. Attention deficit hyperactivity disorder (ADHD) was diagnosed for a small number of individuals (2/28; 1 diagnosed in adulthood). Hyperactivity (2/28), attention difficulties (2/28) and restricted interests and behaviour (2/26) were also noted in further participants without formal ASD or ADHD diagnoses. Mental health conditions such as anxiety (6/28), depression (7/28), and obsessive-compulsive disorder (3/28) were reported in six adults and one adolescent..

Most participants (school aged or older) with pathogenic *FOXP2* variants attended mainstream schools (15/24); seven attended special education schools and two attended a mix of special education and mainstream schools. Learning support (e.g., teaching aide, individualised learning plan) was common (15/24) across all settings. All five preschool participants attended specialist pre-school settings for children with additional learning needs. All caregivers of school-aged children and adolescents reported that their child’s academic progress had been most impacted by their speech and language impairments.

#### Communication development

Speech development was characterised by limited babbling and a reduced phonetic (sound) inventory relative to peers across the first seven years of life, when a full inventory is typically acquired. Some developed first words around the typical age of development (12-15 months, 10/26), whereas others were slightly (15-18 months, 1/26) or more significantly (>18 months, 14/26) delayed (Table 3). One participant (ID 14) had not said their first words yet in early childhood (2-5 years old). Six participants had not yet mastered combining words (IDs 1a, 6, 9, 12a, 13, 15, 18). Only four participants combined words in line with the typical development milestone of 2-3 years of age. Remaining participants combined words between 4 to 5 years (4/22), 6 to 7 years (2/22) and at 8 years or older (5/22), representing protracted development relative to the typical developmental milestone of 2-3 years.

**Table 3.**
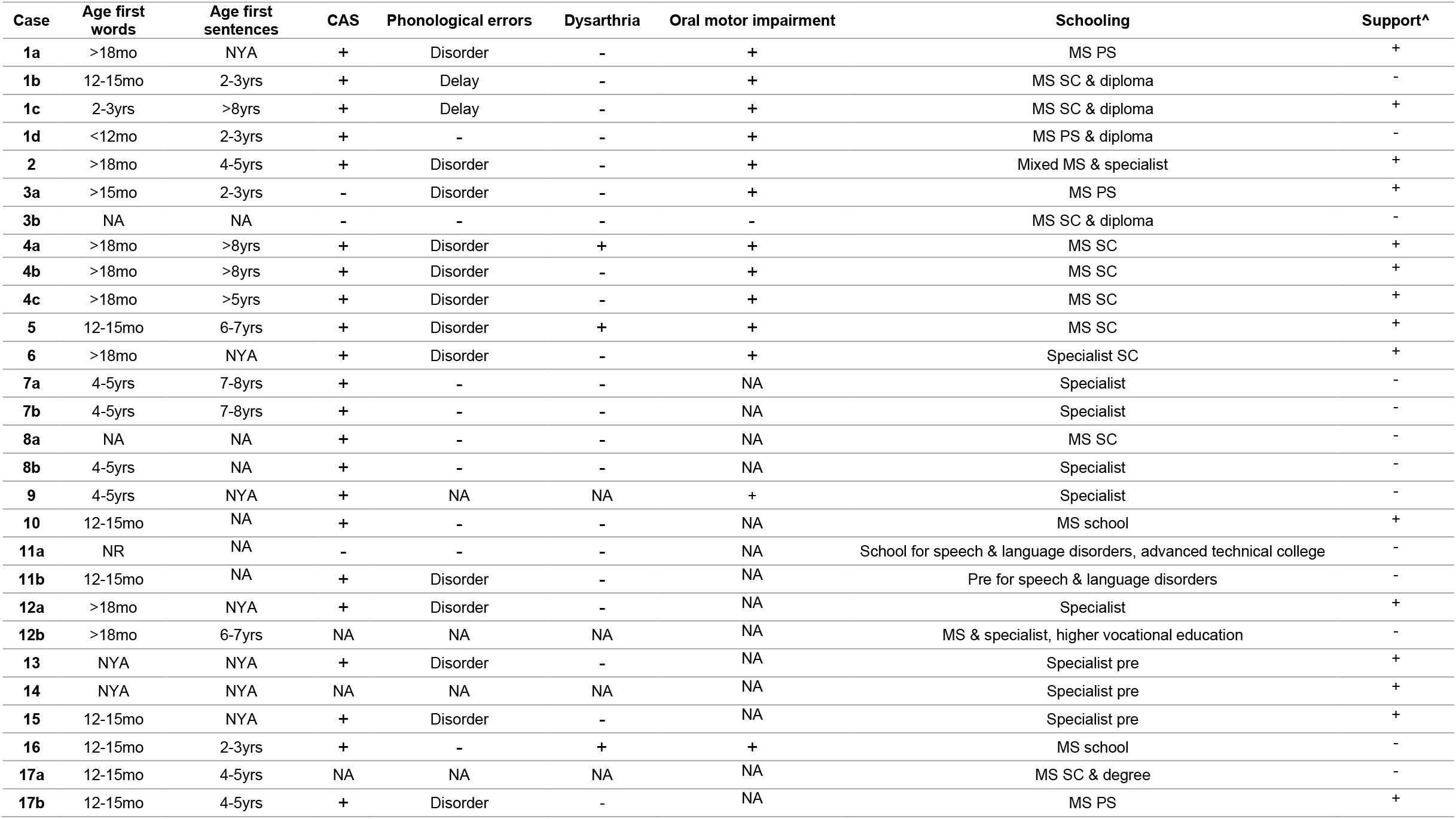
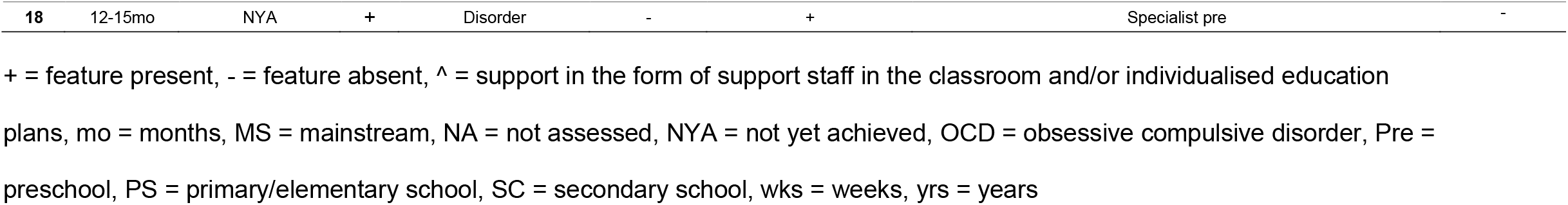
Speech features and educational placement of individuals with pathogenic SNVs/indels disrupting *FOXP2*

### Speech

CAS was the most common speech diagnosis (88.5%, 23/26) (Figure 2, Table 3), with features including frequent sound omissions, the same consonant or vowel being produced differently across different words, impaired sequencing of phonemes and syllables, voicing errors, syllable segregation, difficulty achieving initial articulatory configurations, equal stress, altered suprasegmental features and slow rate (Supplementary Table 3). CAS was present in most English (87.5%, 14/16) and German (87.5%, 7/8) speakers. Participant 16 also had persistent sound repetitions presenting as a mild stutter.

**Figure 2.**
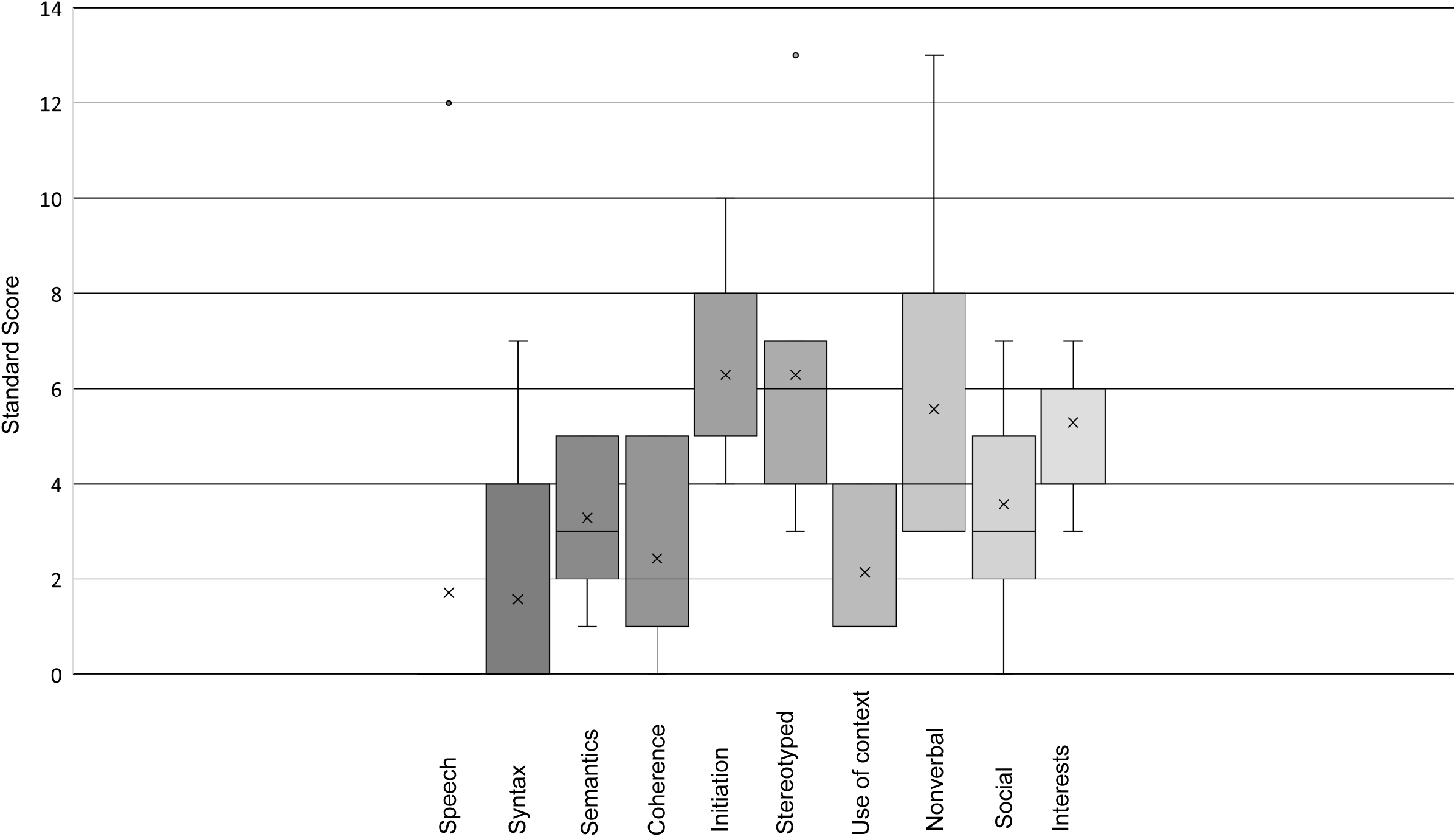

Some participants had multiple co-occuring speech sound disorders (Figure 2). Dysarthria (12%, 3/25) was infrequent and mild in severity, characterised as monoloud and monopitch (ID 16), mixed nasality and harsh vocal quality (IDs 4a, 5). Phonological disorder was common (56%, 14/25), especially in children (<18 years); although for some (IDs 4a, 4b) this persisted into adulthood (Table 3). A further two adults had severe phonological delay, that is, typical phonological error patterns that appear in the speech of younger individuals but that should have resolved by 7 years of age (Supplementary Table 4a, IDs 1b, 1c). Disordered oral motor movements were present both on speech (e.g., say ‘pataka’) and non-speech tasks (e.g., bite then blow) (93%,14/15).

Two adults and one child did not have signs of CAS at time of testing (IDs 3a, 3b, 11a). Participant 11a had a speech and language disorder in childhood for which they received therapy, but could not recall having CAS. The other adult participant (ID 3b) reported being a ‘quiet’ child but did not have speech therapy, and his son (ID 3a) had a phonological disorder without CAS.

Phonetic inventories were analysed for 13 English-speaking participants (Supplementary Table 4a and 4b). Strikingly, 61.5% (8/13) of English-speaking participants did not have the affricate 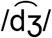 (e.g. ‘j’ in ‘jump’) and 53.8% (7/13) did not have its voiceless counterpart, 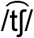 (e.g. ‘ch’ in ‘chair’). Many were also missing the later developing sounds of /ɹ/ (e.g. ‘r’ in ‘rabbit’, 61.5%, 8/13), /θ/ (e.g. voiceless ‘th’ in ‘thin’, 53.8%, 7/13) and /ð/ (e.g. voiced ‘th’ in ‘this’, 53.8%, 7/13) (Supplementary Table 4a and 4b). Other phonemes absent in some English speakers’ inventories were /∫/ (‘sh’, 38.5%, 5/13), /ŋ/ (‘ng’, 38.5%, 5/13), /l/ (25%, 3/12) and /s/ (25%, 3/12). The phonemes 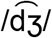 and 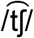 were not present in most German participants (4/6).

Average intelligibility for children, assessed via the ICS, ranged from ‘never’ understood (20%, 2/10), to ‘rarely’ (33.3%, 3/10) to ‘sometimes’ understood (50%, 5/10). For adults, average intelligibility ranged from ‘sometimes’ (28.6%, 2/7) to ‘usually’ understood (57.1%, 4/7). Only one participant was ‘always’ understood (ID 3b).

### Language and literacy

More than half the cohort (57.7%, 15/26) had mild to severe receptive vocabulary impairment (Table 3, Supplementary Figure 2). Receptive grammar was commonly affected (72.2%, 13/18) ranging from moderate to severely impaired. Expressive vocabulary (83.3%, 15/18) and expressive grammar impairments were also common (86.7%, 13/15).

Speech was the most severely affected communication domain (mean score=1.7) for the CCC-2 (completed for IDs 1a, 2, 3a, 5, 6, 15, 17b; Figure 3, normative mean=10, SD=3).

Five participants were minimally verbal (<30 words; IDs 9, 12a, 13, 14, 15; between 2 and 18 years old). Three of the five used high-tech Augmentative and Alternative Communication (AAC) devices (e.g., a speech generating application on a tablet) whilst the remaining two used some sign language.

Spelling (18/25) and reading (18/25) impairments were common (Table 3). This was reflected in the results from the VABS-3 (written subdomain mean=9.25, normative mean=15, SD=2). Four participants were at the pre-literacy stage (<5 years of age.

### Adaptive behaviour and cognition

Receptive and expressive language performance was low in those assessed (n=11, mean=9.5 in both domains, Supplementary Figure 3) and not significantly different between the two domains (p=0.79). Language performance (mean=70.73) was significantly different from (p=0.01) and higher than socialisation (mean=61.18) and daily living (mean=57.45). Motor skills were an area of relative strength, albeit impaired compared to norms (mean=77.18), although the sample size was small and results should be interpreted with caution. Further, normative data for motor skills is only available up to 9 years 11 months, however none of the older participants reached ceiling on motor skills.

Language skills were incongruent with intellectual skills for many, as seven participants with severe language impairment scored within the average range on non-verbal subtests (Table 3). IQ was formally assessed in 21 participants (FSIQ n=10, non-verbal IQ n=11). In the non-verbal testing eight performed in the average range, one was borderline and two were moderately impaired. For a FSIQ, two performed within the average range, four in the borderline range (70-85 IQ) and four had a mild intellectual disability (50-69 IQ).

## Discussion

We systematically delineated the speech and cognitive phenotype in 29 probands from 18 unrelated families (11 of which are novel) with heterozygous pathogenic missense/loss-of-function variants disrupting *FOXP2*, and completed the first cross-linguistic analysis of this disorder. Our data confirm aberrant speech and language development as a central feature. Whilst speech presentation improves over time, with a reduction in CAS severity and improvement in phonological production, the disorder is characterized by impaired speech intelligibility that persists into adulthood, with most adults in our cohort being understood only ‘sometimes’ or ‘usually’ (rather than ‘always’) understood.

In terms of intellectual ability, scores ranged from below average to average in our cohort. For individuals with FSIQ data available most (8/10) were scored as having borderline or mild intellectual disability, while for those who had only non-verbal IQ data available most (8/11) were average. This range and profile is in line with previous findings for individuals with pathogenic SNVs/indels in *FOXP2*, with most individuals falling in the low average range and below for FSIQ and non-verbal IQ. The critical point of note here is that FSIQ takes into account the language metric of vocabulary knowledge, hence why FSIQ is generally more impaired than non-verbal IQ skills for individuals with this speech and language phenotype. At the same time, we also confirm observations from prior literature that the profile of this disorder differs from classic intellectual disability syndromes, in that severe speech and/or language impairments can occur against a background of non-verbal cognition within the normal range (46) as observed for seven of our probands with available data.

ASD has previously been reported in only a small number of individuals carrying pathogenic SNVs/indels of *FOXP2* [n=2/46, Supplementary Table 1]. The findings in our cohort indicate that there may be a higher prevalence of ASD in this disorder (25% of our cohort), than in the general population (∼1-2%) (47), although further research is needed to account for the discrepancy between our current findings and the prior literature. Of note, pathogenic variants in the closely related orthologue *FOXP1* are known to substantially the increase risk for ASD (48). Common non-coding poymorphisms in introns of *FOXP2* have shown associations with ADHD in large-scale genome-wide association studies, in the context of a multifactorial framework (49). The current study clearly shows that, by contrast, high-penetrance SNVs/indels disrupting this same locus do not yield elevated susceptibility to ADHD, with a prevalence in our cohort (2/28 = 7%) that is similar to that in the general population (50).

Sleep disturbances were common in our cohort and have been previously associated with idiopathic CAS (51) and other neurodevelopmental disorders such as ASD, intellectual disability and ADHD (52). The aetiology of sleep problems in such disorders is currently unknown, but they are posited to have biological and psychopathological causes. Although ASD, intellectual disability and ADHD were present here to varying degrees (as discussed above), within our cohort, sleep disturbance is also noted in children without those diagnoses.

We provide novel insights into other clinical diagnoses of mental health conditions that might be associated with pathogenic *FOXP2* variants. In particular, anxiety (21%) and depression (25%) had a higher prevalence than in the general population (between 2-4%) (50) and than in other neurodevelopmental disorders which were also present in our cohort, such as mild intellectual disability (∼3-4%) (53). Anxiety has previously been associated with idiopathic CAS (54, 55), and speech and language disorders are known to have possible negative impacts on mental health (54-57). It is difficult to ascertain whether mental health disorders are part of the phenotypic spectrum due to pathogenic *FOXP2* variants, or occur as a secondary consequence of the communication deficits experienced by affected individuals, as is seen in other speech disorders, such as stuttering (58). All participants with anxiety and depression were older than 16 years old, perhaps indicating that these mental health conditions arise later in life due to the impact of the communication impairment.

Gross motor impairment is thought to be relatively uncommon in individuals with pathogenic SNVs/indels disrupting *FOXP2* (16). However, two-thirds of the assessed participants in this study indicated having difficulty with gross motor skills during development. *FOXP2* disrupton therefore appears to impact brain circuits involved in fine as well as gross motor development. Gross motor skill learning deficits have been identified in knock-out animal models (59).

We did not find convincing evidence of a dysmorphism phenotype in individuals with pathogenic *FOXP2* variants. Although physical features were noted for 82% of participants, most were minor and only shared within individuals from the same family. There was no consistent pattern of morphology seen across multiple unrelated probands in the cohort.

Regardless of the associated developmental features noted here, CAS was the most striking and consistent phenotypic characteristic in the present cohort. Dysarthria was far less common than CAS, clarifying the role of *FOXP2* in the planning and programming of movement sequences, as supported by animal models (59). Adult participants typically had more intact, albeit incomplete, phonetic inventories than younger participants. In our study, more than half of all English-speaking participants with pathogenic missense/loss-of-function variants in *FOXP2* were missing one or more of the phonemes: 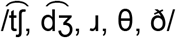 from their inventory. Three of these phonemes (/ɹ, θ, ð/) are in the ‘late eight’ sounds of English speech development, whilst affricates 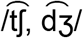 sit within the ‘middle eight’, referring to whether they are acquired earlier or later during typical phoneme acquisition (60).

There may be a window for plasticity and acquisition of new phonemes. Children with speech sound disorders are more likely to have persistent speech error patterns if these are not resolved by 8-and-a-half years old (60). We might speculate that *FOXP2* dysfunction has greatest impact between 2 and 7 years old when most phonemes are acquired (61, 62). Intriguingly, neural expression of orthologues of *FOXP2* in model organisms has been shown to vary during different periods of vocal development (63-65), for example being upregulated in parts of the brain of the male Zebra finch during a developmental window that is important for vocal learning (66). Reduced expression of this gene in mice alters development and continuing plasticity of neuronal networks (67), impairs synaptic plasticity in striatial and cerebrellar circuits, and affects learning of motor skills (68, 69). Perhaps the lack of acquisition of ‘late eight’ sounds and affricates in children with *FOXP2* disruptions may relate to the closing of the relevant developmental window. Other theories to explain the lack of acquisition of these phonemes include reduced functional load and ambient frequency for these phonemes (70) or the motoric complexity of these sounds (73) which rely heavily on tongue co-ordination and movement, known to be impaired in children with CAS (71-73). Further research is required to disentangle these relationships.

Speech impairment in the three participants without CAS was minimal or even absent, contrasting with unaffected, previously reported cases of *FOXP2* disruption (13). The speech of participant 18 was also less impaired than that of other children, although he had a diagnosis of CAS. Participant 11a was not referred for genetic testing for his speech, but rather to determine whether the variant in his child was *de novo* or inherited. Of note, family 3 and participant 18 were the only participants who had not been referred for testing primarily on the basis of speech and learning impairments. Thus, it is possible that there is a broader range of speech phenotypes associated with pathogenic *FOXP2* variants, and that individuals with milder presentations are unlikely to be referred for genetic testing.

Although there was no statistically significant difference between receptive and expressive language as reported by caregivers, other standardised tests indicated receptive language was more intact than expressive language. Literacy skills were also low across the cohort, in line with the high rate of literacy challenges for individuals with idiopathic CAS (74).

The speech domain on the CCC-2 confirmed that speech was the most severely impaired form of communication in children. AAC systems should be considered for children with pathogenic *FOXP2* variants due to the protracted speech development and severe speech impairment which persists for many throughout their lifetime.

We were unable to identify any clear phenotype-genotype correlation in the present cohort as we did not have sufficient power due to too few cases of missense and loss-of-function variants. The severity of speech and language disorder differed even amongst individuals with the same *FOXP2* variant in the same family. Family 3 had a relatively mild presentation compared to the other individuals of the study, despite having an intragenic deletion encompassing all *FOXP2* exons. Participant 18 had a translational start-site variant and a mild phenotype, with a larger phonemic repetoire and expressive vocabulary than other participants of similar ages in the cohort. This variant may not cause a clearcut loss-of-function since there are alternative transcription start-sites, potentially leading to a shorter protein.

In conclusion, CAS and language impairments are the most discernable features associated with heterozygous pathogenic missense/loss-of-function variants disrupting *FOXP2*. We also provide evidence of additional neurodevelopmental features in subsets of our cohort, such as mild intellectual disability, ASD, anxiety, depression, and sleep disturbances. There appears to be no distinctive physical features consistently associated with *FOXP2* disruptions. The phenotype associated with pathogenic variants that directly disrupt *FOXP2* remains relatively specific to speech disorder, compared to phenotypes associated with other monogenic conditions involving CAS (74). Thus, our findings demonstrate that *FOXP2* provides an especially valuable entrypoint for examining the neurobiological bases of speech disorder.

## Data Availability

The datasets generated and analysed during this study are not publicly available because participants have not given permission for data to be made public but may be requested from the corresponding author (ATM) who could go back to the participants to request data sharing. Genotypic and phenotypic data were submitted to Decipher.

https://decipher.sanger.ac.uk/

## Acknowledgements

Sincere thanks to the children, families and clinicians who took part in this project. Several authors of this publication are members of the European Reference Network on congenital malformations and rare intellectual disability (ITHACA).

## Ethical Approval and Consent

Ethics approval was obtained from the Royal Children’s Hospital, Melbourne, Human Research Ethics Committee (HREC 37353A). Adult participants and caregivers of child participants provided informed consent to participate in the study and for results of this study to be published.

## Data Sharing

The datasets generated and analysed during this study are not publicly available because participants have not given permission for data to be made public but may be requested from the corresponding author (ATM) who could go back to the participants to request data sharing. Genotypic and phenotypic data were submitted to Decipher (https://decipher.sanger.ac.uk/).

## Author Contributions

**LDM:** generated data, analysed data, interpreted data, wrote manuscript

**EM:** generated data, analysed data, interpreted data, revised manuscript

**MS:** generated data, analysed data, revised manuscript

**ISW:** generated data, analysed data, revised manuscript

**BV:** generated data, analysed data, revised manuscript

**KS:** generated data, analysed data, revised manuscript

**TB:** generated data, analysed data, revised manuscript

**RB:** generated data, analysed data, interpreted data, revised manuscript

**AV:** generated data, analysed data, interpreted data, revised manuscript

**DTL:** collected data, analysed data, revised manuscript

**CP:** generated data, analysed data, revised manuscript

**EB:** generated data, analysed data, revised manuscript

**ST:** generated data, analysed data, interpreted data, revised manuscript

**UM:** generated data, analysed data, revised manuscript

**AR:** generated data, analysed data, revised manuscript

**FL:** contributed data, revised manuscript

**DK:** generated data, revised manuscript

**DA:** analysed data, interpreted data, revised manuscript

**TK:** generated data, revised manuscript

**SEF:** designed and conceptualised study, directed project, analysed data, interpreted data, wrote manuscript

**CZ:** designed and conceptualised study, directed project, generated data, analysed data, interpreted data, revised manuscript

**ATM:** designed and conceptualised study, directed project, generated data, analysed data, interpreted data, wrote manuscript

