## Supplementary Material Table 1 for "Indepth characterization of a cohort of individuals with missense and loss-of-function variants disrupting *FOXP2*"

**Supplementary Table 1** Cases of *FOXP2*-related disorder previously published in the literature ordered reverse chronologically

| Author | Variant | N | Age | Sex | Speech | Oromotor | Language | Cognition | Physical features | Motor | Other | FOXP2-only or plus |
|---|---|---|---|---|---|---|---|---|---|---|---|---|
| Rieger *et al* 2020 | 4.7 Mb deletion arr[GRCh37] 7q31.2q31.31 (downstream of *FOXP2*) | 3 | 10 | M | Spoke in full sentences with impaired articulation, delayed speech development | NR | Below average language, delayed language development | Hyperactivity, attention difficulties | Mild dysmorphic features | Motor milestones typical | MRI NAD | Plus |
| | | | NR *de novo* | F | Dysarthria | NR | | | | | Attended special school | Plus |
| | | | 3y3m | F | Far below average for speech production, speech delay, imprecise articulation | NR | Normal comprehension of sentences, only 25-word vocabulary, age-appropriate development on first words | | Mild dysmorphic features | NR | | Plus |
| Argyropoulos *et al* 2019 | missense (p.Arg553His ) **KE FAMILY** | 10 | | | Poor performance in non-word repetition | Poor performance non-speech oromotor tasks | | | | | *f*MRI: pronounced volume reduction right hemispheric & total cerebellar Crus I | Only |

| | | | | | | | | | | | |
|---|---|---|---|---|---|---|---|---|---|---|---|
| Akahoshi & Yamaoto 2018 | *de novo* intersitial deletion 7q31.1q31.3 | 1 | 29y | F | NR | Hypotonia & poor sucking in infancy | NR | Binet–tanaka test: 53 (mild ID) | Horizontal line in her palm, high-arched palate, mandibular protrusion | Gross motor delayed | Sleep difficulties, schizophrenia | Plus |
| Reuter *et al*, 2017 | *de novo* 14kb deletion 7q31.1 | 2 | 11y | F | Short sentences with slurred articulation | NR | Reading & spelling below average | FSIQ 78 VCI 77, PRI 86, WMI 82, PSI 88 | Mild dysmorphic features | NR | Delayed, EEG NAD | Only |
| | | | 11y | F | Short sentences with slurred articulation | NR | Reading & spelling below average | FSIQ 79 VCI 67, PRI 88, WMI 82, PSI 103 | Mild dysmorphic features | NR | Delayed, EEG NAD | Only |
| Reuter *et al*, 2017 | nonsense (p.Arg353*) | 2 | 14y | M | Dyspraxia, slurred articulation | NR | Severely impaired expressive & receptive | FSIQ 72 VCI 57, PRI, WMI, PSI 77-96 | Mild dysmorphic features | NR | Delayed, EEG NAD | Only |
| | | | 35y | F | Dyspraxia, slurred articulation, stuttering | NR | NR | Learning difficulties (self-reported) | NR | NR | | Only |
| Reuter *et al*, 2017 | missense (p.Arg561Pro) | 2 | 10y10m | F | Single words only | NR | Severely impaired | FSIQ 70 | Mild hypotonia, no dysmorphic features | Delayed | Autistic features, pneumonia | Only |
| | | | NR | M | Delayed speech development | NR | NR | NR | NR | NR | NR | Only |

| | | | | | | | | | | | |
|---|---|---|---|---|---|---|---|---|---|---|---|
| Reuter et al, 2017 | nonsense (p.Arg503*) | 1 | 6y | F | Persistent speech impairment, delayed speech development | Choking in infancy | NR | Average IQ, poor auditory memory | Mild dysmorphic features | NR | Strabismus, hyperopia, astigmatism possible seizure at 10m | Only |
| Reuter et al, 2017 | *de novo* nonsense (p.Arg589*) | 1 | 9y11m | M | Vocabulary of 2 words | NR | Impaired expressive & receptive | NR | Mild dysmorphic features | Impaired fine motor skills | Delayed, ASD, left exotropia | Only |
| Reuter *et al*, 2017 | *de novo* frameshift (p.Phe563Leufs*28) | 1 | 19y | M | Stuttering, slurred articulation, stilted intonation, delayed speech development | NR | NR | Good verbal comprehension (not formally tested) | Mild dysmorphic features, 6cm café au lait macule | NR | ASD | Only |
| Reuter *et al*, 2017 | nonsense (p.Arg503*) | 3 | 6y6m | M | Vocabulary of 5 words, delayed speech development | NR | NR | NR | Mild dysmorphic features | NR | Hyperactivity, tantrums | Only |
| | | | 4y6m | F | Three-word sentences | NR | NR | NR | Mild dysmorphic features | Walking at 18m due to hip dysplasia | | Only |
| | | | 36y | M | Simple sentences with slurred pronunciation | NR | NR | NR | Mild dysmorphic features | Delayed | | Only |
| Reuter *et al*, 2017 | missense (p.Pro530Leu) | 2 | 6y6m | M | Slurred articulation, unintelligible, delayed speech development | NR | Average vocabular, below average receptive grammar | FSIQ 72 | NAD | NR | | Only |

| | | | | | | | | | | | | |
|---|---|---|---|---|---|---|---|---|---|---|---|---|
| | | | 40y | M | Articulation disorder, delayed speech development | NR | NR | Average | NR | NR | Strabismus, right exotropia MRI NAD, | |
| Turner *et al*, 2013 | *de novo* frameshift p.Gln415Val*5 | 1 | 8y | M | CAS & dysarthria | Oral motor dyspraxia, mild oral dysphagia | Severely impaired expressive & receptive severe reading & spelling impairment | FSIQ 73 VCI 71, PRI 94, WMI 77, PSI 70 | Submucous cleft palate (repaired) | Motor apraxia | Delayed, strabismus, MRI & EEG NAD | Only |
| Zilina *et al*, 2012 | 8.3Mb deletion 7q31.1-q31.31 | 2 | 3yr | F | CAS | Difficulty chewing, swallowing, drooling | Poor vocabulary | Moderate developmental delay | Dysmorphic | NR | Delayed, autistic features, kidney & eye abnormalities | Plus |
| | | | 28y | F | CAS | Difficulty chewing, swallowing, drooling | NR | Low average FSIQ 88 | Dysmorphic | NR | Delayed, poor social skills, eye abnormalities | Plus |
| Zilina *et al*, 2012 | 6.5Mb deletion 7q31.1-q31.2 | 2 | 6y | F | CAS | NAD | Poor vocabulary | Moderate ID | Dysmorphic, mild ataxia | Motor delayed | Aggressive | Plus |
| | | | NR | F | CAS | NAD | NR | Apparent ID | NR | Motor delayed | Aggressive | Plus |
| Palka *et al*, 2012 | *de novo* 14.8Mb mosaic deletion 7q31.1-q31.3 | 1 | 10y | F | CAS | NR | Impaired expressive & receptive | Borderline FSIQ 71, PIQ, 88, VIQ 57 | High arched palate, lordosis | Fine motor praxis & balance problems | Delayed | Plus |

| | | | | | | | | | | | | |
|---|---|---|---|---|---|---|---|---|---|---|---|---|
| Rice *et al*, 2012 | 1.57Mb deletion 7q31.1-q31.2 | 2 | 4y10 | M | Severe CAS, delayed speech development | Messy bottle feeder, gagging & drooling | Severely impaired expressive, average receptive | Borderline FSIQ 71, PIQ 75, VIQ 77 | NAD | Fine & gross motor planning difficulty | NR | Plus |
| | | | 24y | F | CAS, dysarthria, delayed speech development | Gagging & drooling as infant, delayed swallow | Severely impaired | Low average FSIQ 89, PIQ 92, VIQ 87 | Surgery for L esotropia | Motor NAD | PPD-NOS | Plus |
| Roll *et al*, 2010 | missense (p.Met406Thr) | 4 | NR | 2F 2M | NR | NR | Impaired (proband) | Cognitive impairment (proband) | NR | NR | Polymicrogyria (proband) | Only |
| Tomblin *et al*, 2009 Shriberg *et al*, 2006 | balanced translocation t(7;13) (q31.1;q13.2) disrupting *FOXP2* | 2 | 50-52y | F | CAS & spastic dysarthria | No orofacial apraxia | Impaired expressive & receptive | Low average FSIQ 88, PIQ 95 VIQ 81 | NAD | NR | | Plus |
| | | | 18-20y | F | CAS & spastic dysarthria | No orofacial apraxia | Impaired expressive & receptive | Low average FSIQ 81, PIQ 8D, VIQ 81 | NR | NR | | Plus |
| Lennon *et al*, 2007 | 9.1Mb deletion 7q31.1–7q31.31 | 1 | 7y4m | M | CAS | Drooling, low oral-motor tone | Severely impaired expressive & receptive | Moderate ID | Dysmorphic | | Global delay | Plus |
| Zeesman *et al*, 2006 | *de novo* 16Mb deletion 7q31.2-q32.2 | 1 | 5y | F | Verbal dyspraxia | Oromotor dyspraxia | Severely impaired expressive & receptive | Below average-average | Dysmorphic | | Delayed | Plus |
| Feuk *et al*, 2006 | *de novo* deletion 7q31.1-q31.3 (15Mb Patient 1; 11Mb Patient 3); 7q31.2-q32 | 5 | | NR | CAS | Oromotor difficulties | Impaired expressive & receptive | Cognitive impairment | NR | NR | Delayed, patient 3: ASD | Plus |

| | | | | | | | | | | | | |
|---|---|---|---|---|---|---|---|---|---|---|---|---|
| | (13Mb Patient 2; 15Mb Patient 4); 7q22-q31.3 (15Mb Patient 5) | | | | | | | | | | | |
| Feuk *et al*, 2006 | translocation t(3;7)(q23;q31.2) disrupting *FOXP2* | 1 | NR | NR | CAS | Oromotor difficulties | Impaired expressive & receptive | Cognitive impairment | NR | NR | Delayed | Plus |
| Feuk *et al*, 2006 | *de novo* deletion 7q31.2-q32 (26Mb Patient 18; 14Mb Patient 20); 7q22-q31.33 (22Mb Patient 19); q31.1-q33 (30Mb Patient 21 & 22) | 5 | NR | NR | Severe dyspraxia | NR | Language delay | NR | NR | NR | Patient 18: ASD, global delay (patients 18, 21 & 22) | Plus |
| Feuk *et al*, 2006 | maternal uniparental disomy of chromosome 7 that reduces *FOXP2* expression | 7 | NR | NR | CAS, delayed speech development | Oromotor dyspraxia | Impaired expressive, average receptive | NR | NR | NR | Silver-Russell Syndrome Patient 13: ASD | Plus |

| | | | | | | | | | | | | |
|---|---|---|---|---|---|---|---|---|---|---|---|---|
| MacDermot *et al*, 2005 | nonsense (p.Arg328*) | 3 | 4y | M | CAS | NR | Impaired expressive & receptive | NR | NR | NR | Delayed | Only |
| | | | 2y11 | F | Minimally verbal | Oropharyngeal dyspraxia | Impaired expressive & receptive | NR | NR | Motor dyspraxia | Delayed | Only |
| | | | NR | F | Poor clarity, delayed speech development | NR | Simple grammar | Comprehension difficulties | NR | NR | | Only |
| MacDermot *et al*, 2005 | Heterozygous insertion of CAGCAGCAACAA into exon 5 | 1 | NR | NR | CAS | NR | NR | NR | NR | NR | | Only |
| Vargha-Khadem *et al*, 1995; Lai *et al*, 2001 | missense (p.Arg553His) **KE FAMILY** | 15 | mean 24.4y | | CAS, delayed speech development | Orofacial dyspraxia | Impaired expressive & receptive, reading/spelling impairments | Borderline VIQ (mean 75), average PIQ (mean 86) | NAD | Motor NAD | Impaired performance on phonological loop working memory assessment (Schulze *et al* 2017) | Only |
| Lai *et al*, 2000 | *de novo* balanced translocation t(5;7)(q22;q31.2) disrupting *FOXP2* | 1 | 5.5y | M | Verbal dyspraxia | Severe orofacial dyspraxia | Impaired expressive & receptive | Normal non-verbal skills | NAD | Delayed | | Plus |

ASD = autism spectrum disorder, CAS = childhood apraxia of speech, EEG = electroencephalogram, FSIQ = full scale intelligence quotient, ID = intellectual disability, m = months, MRI = magnetic resonance imaging, NAD = no abnormalities detected, NR = not reported, PIQ = performance intelligence quotient, VIQ = verbal intelligence quotient, y = years

**Note: A further 15 cases with pure interstitial deletions of chromosome 7 are reviewed in Palka *et al*, 2012, however speech and language data were not reported. Zhao *et al*, 2016 published one case of a female child with a *de novo* 3.2 Mb sub microscopic deletion 7q31.2–7q31.31 (downstream of *FOXP2*) and Sangu *et al* , 2017 reported one case of a male child with a *de novo* de novo 1.9-Mb microdeletion in 7q31.33q32.1 (*FOXP2* is not within deletion region)**
