## Supplementary Material Figure 1 Table 3 Table 4a & 4b for "Indepth characterization of a cohort of individuals with missense and loss-of-function variants disrupting *FOXP2*"

**Supplementary Figure 1.** Family pedigrees of inherited cases of pathogenic missense/loss-of-function variants disrupting FOXP2

This figure has been removed to avoid identification of patients

**Supplementary Table 3** Additional health and medical phenotypic features in individuals with pathogenic missense/loss-of-function variants disrupting FOXP2

This table has been removed to avoid identification of patient

**Supplementary Table 4a** Phonemic inventory of English-speaking participants with pathogenic missense/loss-of-function variants disrupting FOXP2

This table has been removed to avoid identification of patient

**Supplementary Table 4b** Phonemic inventory of German-speaking participants with pathogenic missense/loss-of-function variants disrupting FOXP2

This table has been removed to avoid identification of patient
