## Supplementary Table 2 for "Indepth characterization of a cohort of individuals with missense and loss-of-function variants disrupting *FOXP2*"

**Supplementary Table 2** Speech apraxia features in assessed participants with pathogenic missense/loss-of-function variants disrupting *FOXP2*

| **Speech apraxia features^** | **1a** | **1b** | **1c** | **2** | **3a** | **3b** | **4a** | **4b** | **4c** | **5** | **6** | **7a** | **7b** | **8b** | **10** | **11a** | **11b** | **12a[a]** | **13[a]** | **15[a]** | **16** | **18** |
|---|---|---|---|---|---|---|---|---|---|---|---|---|---|---|---|---|---|---|---|---|---|---|
| **(1) Inconsistent errors** | | | | | | | | | | | | | | | | | | | | | | |
| Same C/V different across different words | | | + | + | | | | + | | + | + | + | + | | | | + | NA | NA | + | | + |
| Same word/syllable different on repetitions (percent) | + | | | | | | | | | + | + | | | | | | + | NA | NA | | | + |
| Inconsistency of production* | 68% | NA | NA | NA | NA | NA | NA | NA | NA | 72% | 44% | NA | NA | NA | NA | NA | NA | NA | NA | NA | | NA |
| **(2) Lengthened & disrupted coarticulatory transitions** | | | | | | | | | | | | | | | | | | | | | | |
| Speech motor behaviours, including groping during sound production | | + | + | | | | + | + | + | + | | | | | | | | | | + | + | + |
| Difficulty sequencing phonemes & syllables | + | + | + | | | | | + | + | + | + | | | | | | | + | + | + | | + |
| Voicing errors | | | | | + | | + | + | + | | + | + | | + | | | + | | | + | + | + |
| Errors increase with word length & phonological complexity | | + | + | | | | + | + | + | | + | | | | + | | | | | | | + |
| Syllable segregation | + | | | + | | | + | + | | + | + | + | + | + | | | | | | | | + |
| Difficulty achieving initial articulatory configurations or transitory movement gestures | | + | + | | | | + | | + | + | | + | | + | + | | | | + | | + | |
| Difficulty maintaining syllable integrity | + | | | + | | | | + | + | | | | | | | | | + | + | + | | + |
| Repetitions of sounds & syllables | | | + | | | | | | | | | + | | + | + | + | | | | | + | |
| Epenthesis/intrusive schwa | | | | | | | | | | | + | | | | | | | | | | + | + |
| Metathesis | | | | | | | | | | | + | + | | | | | | | | | | |
| Addition errors | | | | | | | | | | | | | | | | | | | | | | |
| Frequent omissions (>10) | + | | | | | | + | + | + | | + | + | + | + | | | + | + | + | + | | + |
| Prolongation errors | + | | | + | | | + | | + | | | | | | | | | | | | | |
| Nonphonemic productions/distorted substitutions | + | | | + | | | + | | | | + | + | + | + | | | + | | | + | | + |
| Hypernasality/nasal emissions | | + | | | | | | | | + | | | + | + | + | | | NA | + | + | + | |
| Slowed & disrupted DDK sequence | + | + | | + | + | | | | | + | + | + | | | + | | + | NA | NA | NA | + | + |
| **(3) Inappropriate prosody** | | | | | | | | | | | | | | | | | | | | | | |

| | | | | | | | | | | | | | | | | | | |
|---|---|---|---|---|---|---|---|---|---|---|---|---|---|---|---|---|---|---|
| Equal stress or lexical stress errors | + | | | + | + | + | + | + | + | | | | | + | | + | + | + |
| Altered suprasegmentals | + | | | + | + | + | + | + | + | + | + | + | | | | | + | + |
| Prolongation errors | + | | | + | + | | + | | | | | | | | | | | |
| Slow rate | | + | + | | | + | + | + | + | + | + | | + | NA | NA | NA | + | + |

^ Rated perceptually using the criteria for rating Childhood Apraxia of Speech from the ASHA CAS Technical Report (2007), *Percent of single words said differently over 3 trials, using the DEAP inconsistency subtest (Dodd et al., 2002), + = feature present, NA = not assessed, [a] = Limited assessment of CAS features possible due to being minimally verbal.
