## Supplementary Material Figure 2 for "Indepth characterization of a cohort of individuals with missense and loss-of-function variants disrupting *FOXP2*"

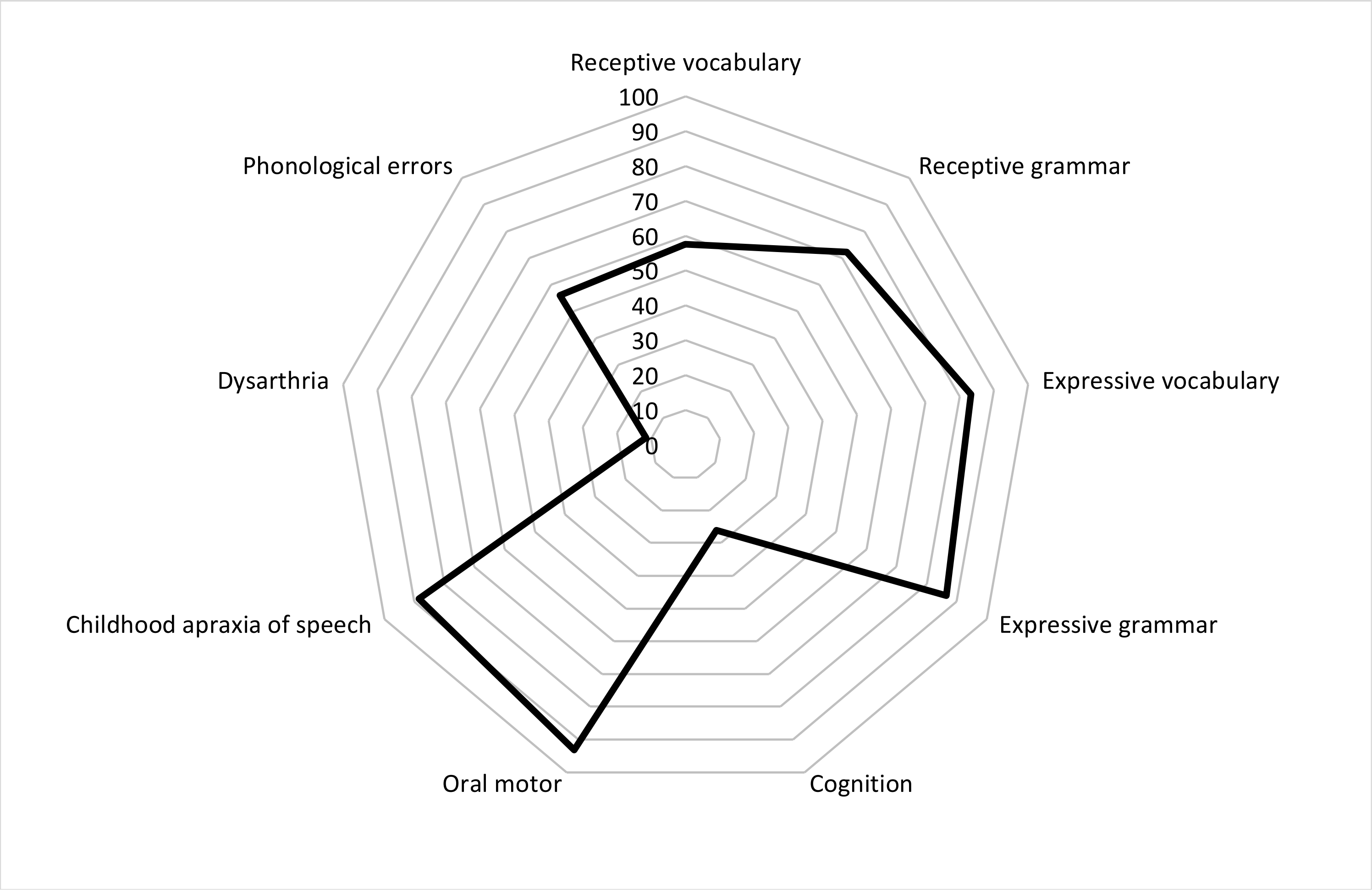


**Supplementary Figure 2.** Phenotypic distribution of speech, language, and intellect in assessed participants with pathogenic *FOXP2* variants.

Cognition refers to general cognitive abilities measured by Full Scale Intelligence Quotient, and perceptual reasoning indices and matrix reasoning sub-tests. Data retrieved from Table 3.
