## Supplementary Material Figure 3 for "Indepth characterization of a cohort of individuals with missense and loss-of-function variants disrupting *FOXP2*"

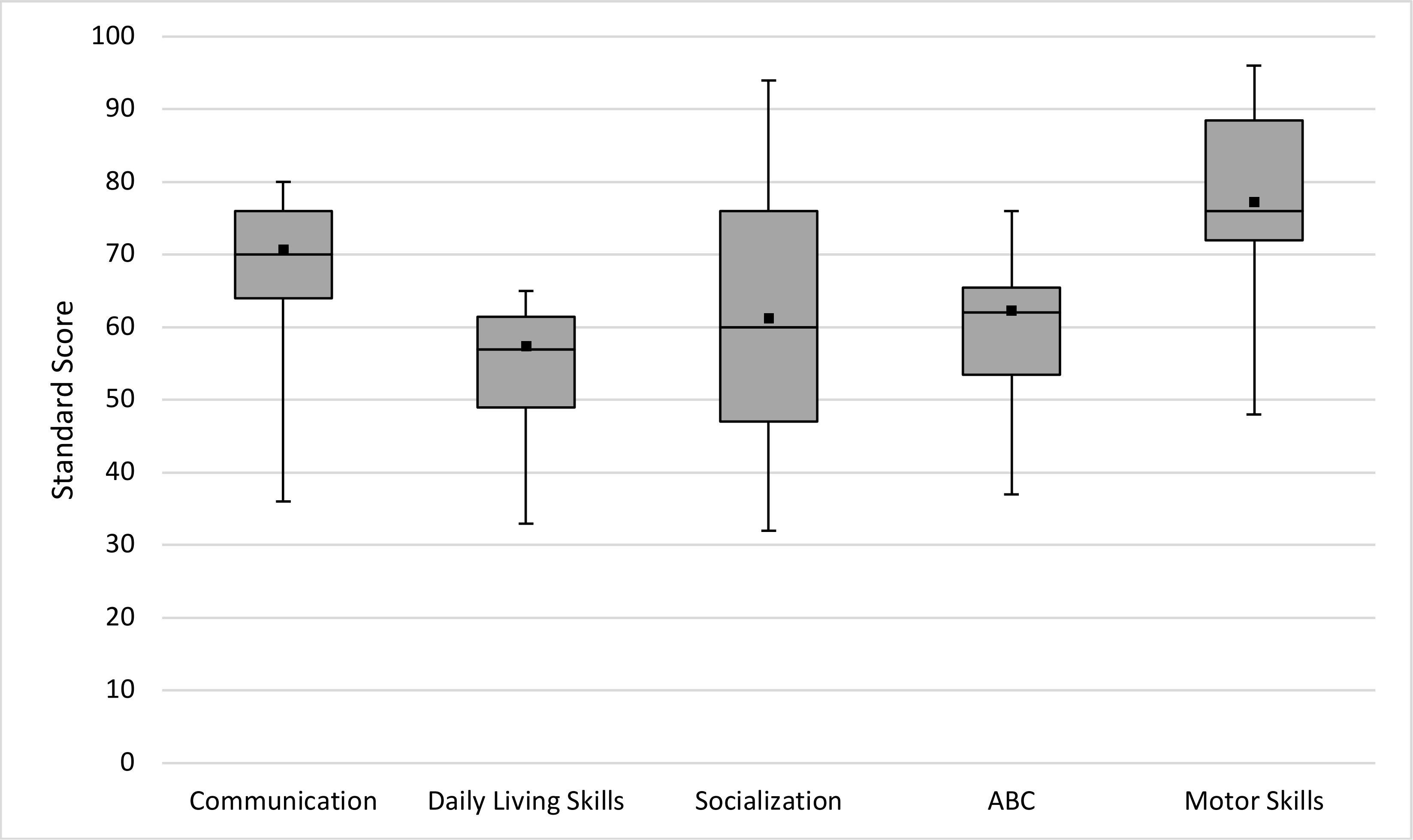


**Supplementary Figure 3 .** Vineland Adaptive Behaviour Scales, Third Edition (42) domains for participants with pathogenic *FOXP2* variants (n=10). ABC = adaptive behaviour composite (overall score). Scores <70 are low, 71-85 moderately low and 86-114 adequate. Line = median, ■ = average.
